# Evaluating mainstreaming in pediatric immunology: an optimal model of care

**DOI:** 10.64898/2026.02.24.26347043

**Authors:** Emily DeBortoli, Tenielle Clinch, Larissa Vaz-Gonçalves, Lucy Burbury, Cian Thomas, Meg Jeppesen, Alberto Pinzon-Charry, Mariana Melo, Anna Sullivan, Matthew F Hunter, Jane Peake, Aideen McInerney-Leo, Peter McNaughton, Tatiane Yanes

## Abstract

**Purpose:** While genomic testing is integral to pediatric inborn errors of immunity (IEI) care, few studies have examined strategies to support its optimal delivery. This study aimed to characterize a pediatric IEI cohort and assess the impact of implementing a mainstream model-of-care (MoC).

**Materials/Methods:** Comprehensive chart audit was conducted for patients (≤18y) who received IEI genomic testing in Queensland, Australia, from 2017-2025. Descriptive analyses captured demographic and clinical characteristics, genomic testing and results, and management outcomes. Inferential analyses assessed changes in genomic practices pre-MoC (<2021) and post-MoC (≥2021).

**Results:** 322 patients met eligibility criteria (n=481 genomic test). Diagnostic yield (27.6%) varied by testing indication, with the highest rate among phagocytic defects (n=4/4;100%) and severe combined immunodeficiency (n=8/10;80%). Very-early-onset inflammatory bowel disease had the lowest diagnostic yield (n=3/68;4.4%), prompting changes to testing criteria. Molecular diagnosis resulted in management changes for 90.5% patients. Genomic testing was widely used pre-MoC (n=251 genomic tests). All outcomes significantly improved pre-and post-MoC (p<0.05): duplicate testing decreased (13.9% to 0%); variants of uncertain significance reduced (37.7% to 7.1%); informed consent documentation increased (70.5% to 88.4%); and diagnostic yield increased (16.2% to 27.4%).

**Conclusion:** Targeted interventions are needed to support delivery of genomic testing and strengthen service effectiveness.

## Introduction

Inborn errors of immunity (IEI) comprise a heterogeneous group of genetic conditions that affect the immune system, causing increased susceptibility to infections, autoimmunity, autoinflammation, malignancies, and/or allergies.^1^ These conditions most commonly present in the pediatric setting, where they are often associated with serious and life-limiting complications.^2^ While IEI are individually rare, the collective prevalence of such conditions is estimated to be between 1:1000 and 1:5000, posing a significant disease burden.^3^ Genomic testing has become an essential aspect of IEI care, with now more than 500 associated genes characterised.^1^ Establishing a molecular diagnosis for such conditions can enable targeted therapeutic interventions and initiation of curative treatment. IEI treatment modalities can include hematopoietic stem cell transplantation (HSCT), immunoglobulin replacement therapy (IRT), gene therapy, and prophylaxis therapies (e.g., antimicrobials and immunomodulators).^4^ Curative therapies are most effective when delivered early in the disease course prior to the onset of infection and/or organ damage, thus increasing the importance of timely molecular diagnosis.^4^ A molecular diagnosis can also inform reproductive decisions, facilitate identification of at-risk relatives, mitigate psychological distress among families, and reduce related healthcare expenditure.^4,5^

Although the clinical and personal utility of genomic testing in IEI is widely recognized, progress in developing models-of-care (MoC) to support its optimal integration into mainstream immunology care remains limited.^6^ There is a lack of guidelines to support patient identification, consent processes, test selection, result communication, and familial considerations.^7,8^ Similarly, few studies have examined the ethical, legal, and social implications (ELSI) of genomics in IEI, and how these factors influence testing practices.^8,9^ The absence of a dedicated MoC can contribute to inefficiencies in healthcare systems and patient harm, such as through missed opportunities for diagnostic testing, delays in the timely delivery of targeted therapies, and identification of at-risk relatives.^10^

To address such challenges, a genomic MoC was developed and implemented for pediatric IEI at the Queensland Pediatric Immunology and Allergy Service (QPIAS), Queensland Children’s Hospital (QCH), an Australian tertiary pediatric hospital.^11^ The QPIAS provides publicly funded, specialist pediatric immunology healthcare to the Queensland population, constituting an expansive geographical area (>1.7M km), with a population of around 5.1 million.^12^ Key features of this genomic MoC include a i) monthly state-wide multidisciplinary team (MDT) meetings to review genomic testing practices, result interpretation, and clinical and familial implications of molecular findings; ii) a genetic counsellor embedded in the service, iii) dedicated role and funding for an clinical immunologist to specialize in genomics, iv) trio-exome sequencing via virtual panel analysis as a front-line diagnostic test,^13^ v) collaborative engagement with the statewide pathology service (i.e. Pathology Queensland), and strong research collaborations, including with the Clinical Immunogenomics Research Consortium Australasia (CIRCA).^14^

This study is part of the BRIDGE (**B**uilding **R**esources for **I**mmunology **D**iagnostic **GE**nomics) project, which aims to optimize pediatric IEI genomic testing practices by considering the needs of children, families, healthcare providers, and health services. Internationally, several IEI registries have aided in the categorization of genomic data, therapeutic interventions, and clinical outcomes.^15^ However, these registries lack data regarding service delivery, limiting the capacity to inform genomic testing practices. Furthermore, there is no Australian data describing local pediatric IEI cohorts.

Thus, in the context of a real-world clinical setting, this study aims to i) characterize an Australian IEI pediatric cohort and genomic testing practices, and ii) assess the impact of implementing a genomic MoC in this setting and identify areas for service improvement. Ultimately, such findings will inform future clinical guidelines and genomic testing practices, thereby leading to an enhancement in clinical care of pediatric IEI patients.

## Materials and Methods

### Ethics Approval

This study received ethics approval from QCH (HREC/24/CHQ/106207) and the University of Queensland (2024/HE000776) and Queensland Health Public Health Act approval (PHA/106207).

### Patient Eligibility and Data Collection

Individuals were eligible for inclusion if they had IEI genomic testing from 2017 to 2025 and were aged ≤18 years at the time of testing. There was no exclusion criterion based on the modality of genomic testing or testing outcome (i.e., positive, negative, variant of uncertain significance (VUS)). Patients were excluded if the genomic test report could not be located, and therefore, results could not be confirmed. Eligible patients were identified via three pathways i) genomic testing ordered by a QPIAS clinician (list generated by Pathology Queensland), ii) private laboratory online portals for QPIAS clinicians, and iii) review of internal QPIAS files to identify additional avenues of testing and to capture patients who had been transferred to another service for clinical care post-genomic testing.

A genomic testing chart review was completed for all eligible patients by authors T.C., A.S., P.M., and T.Y., with the following data collected: demographic information, instances of genomic testing, genomic testing details, and outcomes of genomic testing (**Supplementary Table 1**). Instances of genomic testing ordered for IEI patients by both immunology and non-immunology specialists were included to reflect real-world practices. Due to the lack of accurate documentation of research participation, identifying patients who underwent research testing required collaboration with the clinicians involved to confirm testing and outcomes. All genomic test data were documented in a RedCap database hosted at QCH. Additionally, author L.B. independently reviewed 10% of entries to ensure data accuracy and assist with complex cases. Any discrepancies between authors were resolved through discussion until consensus was achieved.

### Data Categorization and Analysis

Basic descriptive statistics summarized demographic information, indications for testing, number and/or timing of genomic tests, ordering specialty, test modality, results and patient management outcomes. Geographical remoteness was assessed using the Australian Statistical Geography Standard (ASGS),^16^ and socio-economic status was determined using the Socio-Economic Indexes for Areas (SEIFA),^17^ both calculated via postcode data according to the Australian Bureau of Statistics. The cohort distribution based on the ASGS and SEIFA was compared to the 2021 Queensland census data using chi-square analysis.^12^

Indications for testing were categorized based on the Australasian Society for Allergy and Immunology (ASCIA) guidelines (e.g., autoinflammation, atypical infections, bone marrow failure).^18^ Testing modality categories included chromosomal microarray (CMA), karyotype, single gene, panel, virtual panel, exome sequencing, genome sequencing, familial testing, research testing, and other (e.g., mitochondrial point analysis, fragile X analysis). Virtual panel test information was categorized according to the corresponding Australian PanelApp, which the QPIAS clinicians actively contribute towards.^19^ Positive results were categorized according to the 10 respective International Union of Immunological Societies (IUIS) categories, which are defined by the specific clinical and immunological characteristics.^1^

Diagnostic results were assessed to investigate the impact on management and treatment recommendations. Major management changes included recommendations for HSCT, IRT, other targeted treatment and management, prophylaxis monitoring, or changes to HSCT conditioning regimen. Minor management recommendations included changes to existing monitoring. Patients who underwent multiple genomic tests were reviewed to describe the testing pathway and identify potential redundancies (e.g. duplicate testing). Data related to genomic testing practices were grouped into four stages aligned with the implementation of the genomic MoC:

- <2018: pre-implementation of the MoC
- 2019-20: introduction of point-of-care genomic testing (pre-embedded genetic counsellor)
- 2021-22: implementation and pilot testing of the MoC (including embedded genetic counsellor)
- 2023-25: formal establishment of the MoC

The impact of the MoC on genomic testing practices and clinical outcomes were compared to those pre-MoC (pre-2021) using chi-square analysis. Statistical significance was inferred at p<0.05.

## Results

### Cohort Socio-demographic Characteristics

Of the 362 patients identified, 40 did not meet study eligibility and were excluded (n=31 genomic test could not be verified, n=1 genomic testing initiated >18years of age, and n=7 received only non-IEI related genomic testing). Thus, 322 patients were included in the study, of whom 312 (96.9%) of the cohort were still living at the time of data collection (**Table 1**). The mean age at first genomic testing was 5.9 years (range:0-17 years), and 173 (53.7%) of the cohort were assigned male at birth. Most patients resided in a major city (n=233;72.4%) or inner regional area (n=38;11.8%) as per ASGS. Almost half of the cohort resided in an area classed by SEIFA as socio-economically advantaged: most advantaged (n=57;17.9%) or advantaged (n=90;28.3%). Patient distribution was comparable to the Queensland general population (p>0.05), and 17 patients resided in Northern New South Wales. The most frequent indications for genomic testing were very early onset inflammatory bowel disease (VEO-IBD) (n=68;21.1%), diseases of immune dysregulation (n=36;11.8%), atypical infection (n=32;9.9%), predominantly antibody deficiency (n=32;9.9%), and autoinflammation (n=28;8.7%). The indication for testing was unable to be determined for 45 patients (14.0%) (**Figure 1**). Notably, two patients were first identified via a positive severe combined immunodeficiency (SCID) newborn screening test (implemented in May 2023) and later confirmed by genomic testing.

**Figure 1:**
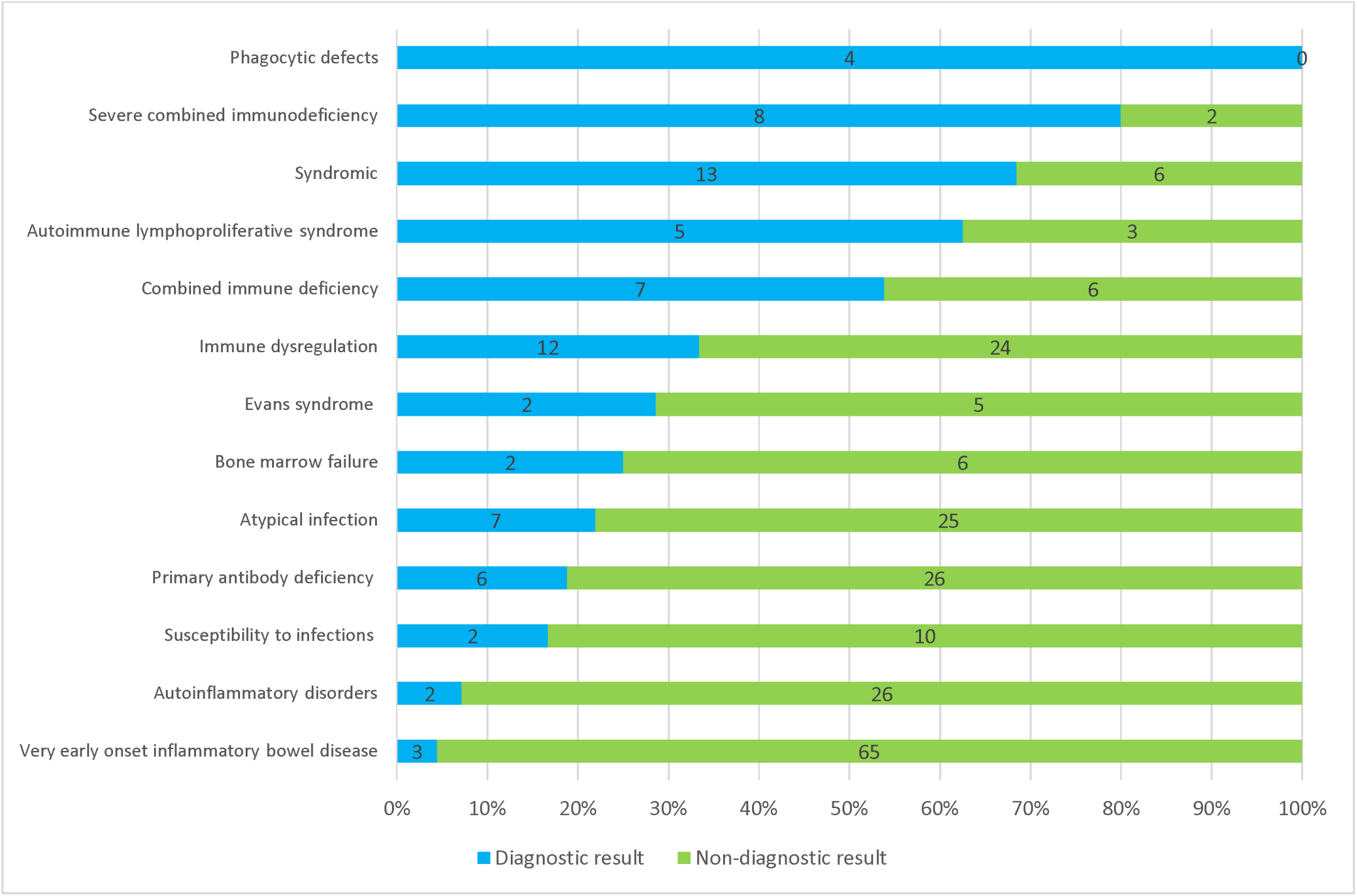
Clinical Indications and Diagnostic Outcomes of Genomic Testing*. *Excludes familial and unknown cases

**Table 1:** Cohort Demographic.

| Characteristic | n (%) |  |
| --- | --- | --- |
| Sex assigned at birth |  |  |
| Female | 149 (46.3) |  |
| Male | 173 (53.7) |  |
| Age at first genomic test (years old) | Mean<br>(range) |  |
|  | 5.9<br>(0-17) |  |
| Living status |  |  |
| Currently living | 312 (96.9) |  |
| Deceased | 10 (3.1) |  |
| Geographical classification (ASGC) | n (%) | Qld population<sup>a</sup> |
| Major city | 233 (72.4) | 3,426,582 (67.0) |
| Inner regional | 38 (11.8) | 920,816 (18.0) |
| Outer regional | 29 (9.0) | 684,865 (13.4) |
| Remote | 1 (0.3) | 33,465 (0.7) |
| Very remote | 4 (1.2) | 46,338 (0.9) |
| Outside of Queensland | 17 (5.3) | NA |
| Socio-economic status (SEIFA) |  |  |
| Most advantaged | 57 (17.9) | 852,726 (16.7) |
| Advantaged | 90 (28.3) | 1,276,043 (24.9) |
| Middle | 74 (23.3) | 1,223,084 (23.9) |
| Disadvantaged | 42 (13.2) | 796,027 (15.6) |
| Most disadvantaged | 42 (13.2) | 964,186 (18.9) |
| Outside of Australia | 17 (5.3) | NA |

<sup>a</sup>Cohort is proportional to the Queensland population (p>0.05, chi-square test). Combined remote and very remote as each p<0.5

### Genomic Testing Practices

A total of 481 genomic tests were ordered for the 322 patients (M:1.5 tests/patient, range:1-5 tests), with 251 genomic tests performed pre-MoC (<2021). More than a third of patients (n=115;35.7%) received ≥2 genomic tests. Most genomic tests (n=249;51.8%) were ordered by QPIAS pediatric immunologists followed by gastroenterologists (n=55;11.4%), geneticists (n=37;7.7%), and rheumatologists (n=21;4.4%) (**Figure 2**).

**Figure 2:**
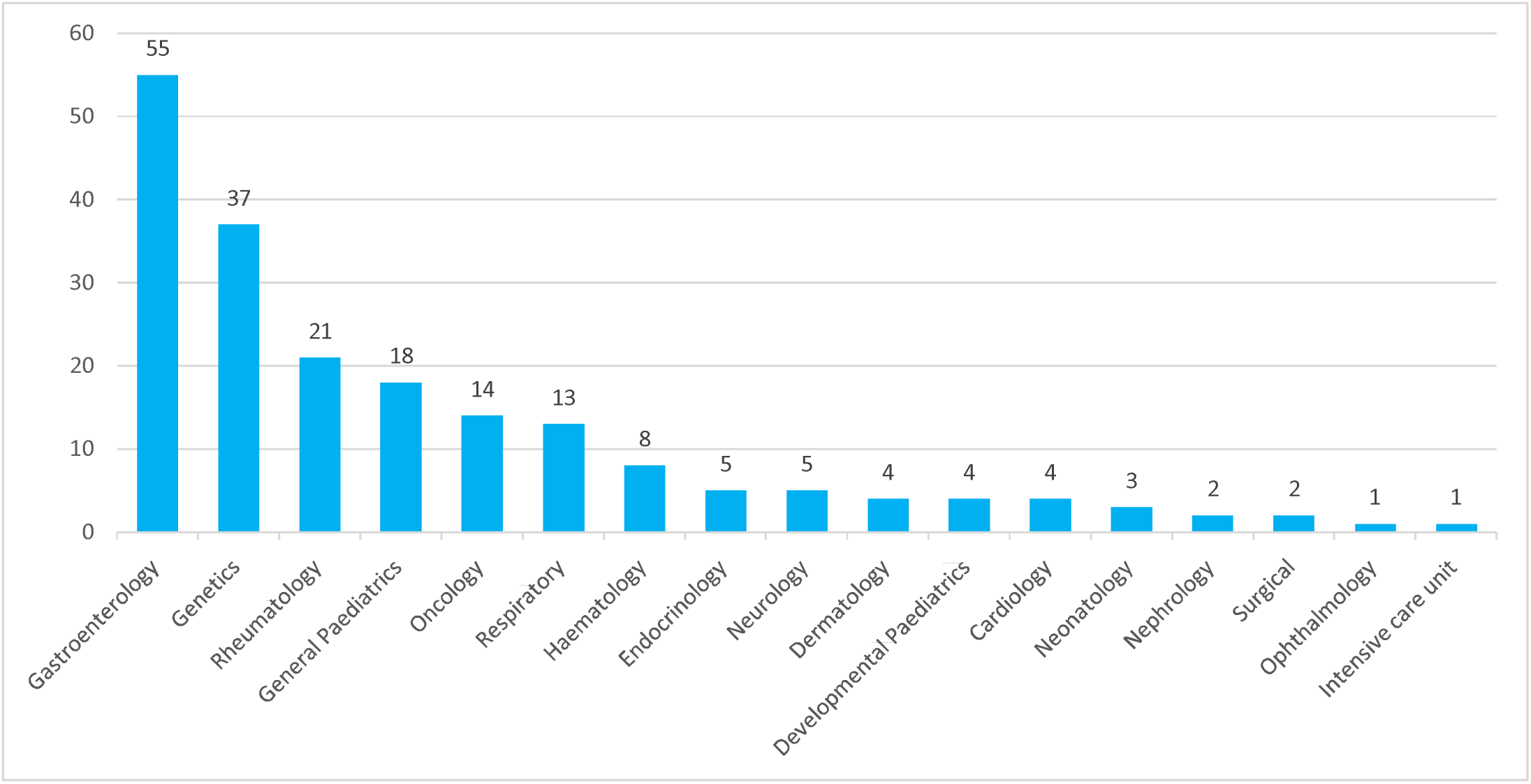
Genomic Testing Ordering Departments*. *Excludes immunology initiated testing and unknown cases

Of the 115 individuals with ≥2 genomic tests, 35 patients had duplicate tests, of which 16 were ordered by different specialties, 16 ordered within the same specialty, and 3 where the ordering specialty was unknown. Examples of duplicate testing included repeat familial testing for the same variant in the same individual (n=1), and duplicate panel tests (n=28) or single gene analysis (n=6), all within a short timeframe (range:1 month to 2 years between tests). All subsequent repeat test requests did not reference the original testing or provide a rationale to justify retesting. Thus, excluding duplicate testing, there were 446 instances of unique genomic testing for the 322 patients, with 84 patients receiving ≥2 unique genomic tests (excluding research and other genomic test types).

Most of the unique genomic tests were arranged for diagnostic purposes (n=425;95.3%), with the remainder pertaining to cascade testing of a pediatric relative such as a sibling or cousin (n=14;3.3%), segregation analysis to assist with VUS classification (n=4;0.9%) or testing to inform HSCT donor eligibility (n=3;0.7%). Excluding research and familial tests, the most ordered genomic tests were panels (n=294/415; 70.8%) (**Table 2**). Of these panels, 126 were virtual panels of singletons (n=74;58.7%), duos (n=2;1.6%), or trios (n=50;39.7%). Trio-testing was increasingly utilized for genomic tests (i.e., virtual panels, exome sequencing, and genome sequencing) over time, with 66.7% (n=52/78) tests analyzed as a trio post formal establishment of the MoC (≥2023), as compared to 18.1% prior (n=13/72).

**Table 2:** Unique Instances of Genomic Testing.

| **Test type (n=415<sup>a</sup>)** | **n (%)** | <sup>a</sup>Exclude<br>s<br>diag<br>nost<br>ic<br>rese<br>arch<br>and<br>fami<br>lial<br>testi<br>ng |
| --- | --- | --- |
| Panel | 169 (40.7) |  |
| Virtual panel (WES backbone) | 125 (30.1) |  |
| Single gene | 57 (13.7) |  |
| Microarray | 31 (7.5) |  |
| Whole exome sequencing (mendeliome analysis) | 11 (2.7) |  |
| Whole genome sequencing | 13 (3.1) |  |
| Other (e.g. mitochondrial, fragile X) | 6 (1.4) |  |
| Karyotype | 1 (0.2) |  |
| Unknown | 2 (0.5) |  |
| **Immunology panel type (n=278<sup>b</sup>)** | **n (%)** | <sup>b</sup>Exclude<br>s<br>non-<br>imm<br>unol<br>ogy<br>pan<br>els;<br>cou<br>nts<br>total<br>pan<br>els<br>orde |
| Immunological disorders super panel | 116 (41.7) |  |
| Inflammatory bowel disease | 58 (20.9) |  |
| Autoinflammatory Disorders | 34 (12.2) |  |
| Disorders of immune dysregulation | 31 (11.2) |  |
| Predominantly Antibody Deficiency | 17 (6.1) |  |
| Combined Immunodeficiency | 9 (3.2) |  |
| Severe combined immune deficiency | 7 (2.5) |  |
| Phagocyte defects | 4 (1.4) |  |
| Complement deficiencies | 4 (1.4) |  |
| Defects of intrinsic and innate immunity | 3 (1.1) |  |
| Susceptibility to Viral Infections | 3 (1.1) |  |
| Chronic granulomatous disease | 2 (0.7) |  |

*red, including cases with multiple panels per test*

Across the 294 panel tests, 342 gene panels were applied (n=48 panel tests had ≥2 gene panels applied), with 278 (81.3%) of these panels specific to immunology conditions (**Table 2**). The most frequently ordered immunology panels were immunological disorders super panel (n=116;41.7%), IBD (n=58;20.9%), and autoinflammatory disorders (n=34;12.2%) **(Table 2)**. Non-immunology panels (n=63) included conditions with symptoms involving other systems, including respiratory (n=13), hematology (n=13), metabolic (n=5), neurology (n=4), nephrology (n=3), dermatology (n=3), orthopedics (n=3), gastroenterology (n=3), pediatrics (n=2), or unknown (n=14).

**Figure 3** shows the genomic testing trajectory for the 84 patients who had ≥2 unique tests, excluding research and other genomic testing (i.e., mtDNA point variant analysis, somatic testing). Most patients had ≥2 panel tests, including 43 in the first instance of testing, 42 in the second instance, and seven in the third instance, all of which were panels capturing symptoms affecting different systems. Few patients had expanded panels (n=16), or re-analysis of a previous panel (n=5). One patient had three instances of research testing, all of which occurred ≤2019 with no diagnosis to date. Ten patients initially underwent panel testing, CMA, or exome sequencing and were subsequently offered clinical genome sequencing. Finally, one patient had two separate instances of familial testing, for two distinct familial variants.

**Figure 3.**
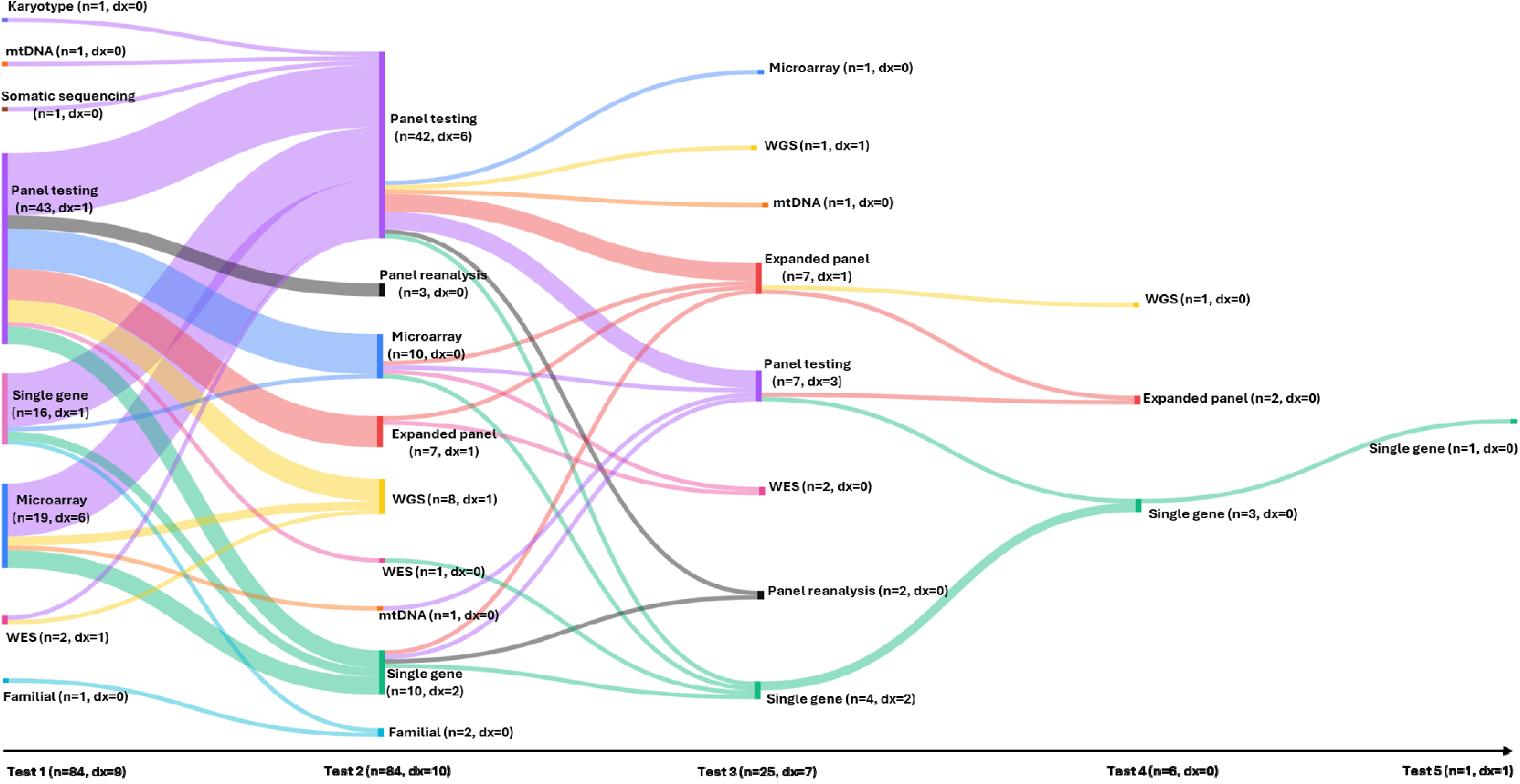
Distribution of Patients Across Genomic Testing Pathways and Diagnostic Outcomes*. Excludes research and other genomic tests. Dx: diagnosis; mtDNA: mitochondrial testing; WGS: Whole Genome Sequencing; WES: Whole Exome Sequencing.

### Diagnostic Results

A diagnostic result was identified for 89/322 patients (27.6%), of which 10 (3.1%) were syndromic cases. Excluding positive results in relatives (n=21), the diagnostic rate for probands was 25.9% (n=78/301). The diagnostic rate was highest among patients who were tested for phagocytic defects (n=4/4;100%), SCID (n=8/10;80%), and syndromic presentations (n=13/19;68.4%) (Figure 2). The lowest diagnostic yield was among patients who were tested for autoinflammatory disorders (n=2/28;7.1%) and VEO-IBD (n=3/68;4.4%). Of those that received a diagnostic result, most patients (n=72/89; 80.9%) received a result in the first instance of genomic testing, and 17 (19.1%) in subsequent testing, which included alternate testing modality (n= 13), the application of a new panel (n=1), expanded panel testing (n=1), or a new single gene analyzed (n=2) (Figure 3). In one case, a diagnostic result was achieved only after five sequential single gene tests, which occurred pre-introduction of the MoC (<2019). Sixteen patients who received a molecular diagnosis from initial testing underwent subsequent genomic testing: eight to investigate clinical features not explained by the original genetic diagnosis, and eight appeared to have duplicate testing with no documented justification for re-testing.

Diagnostic results comprised 77 unique variants in 49 genes and deletions/duplications in five chromosomal regions. Three patients had two unique molecular diagnoses that included a chromosome deletion/duplication with a biallelic pathogenic variant in an IEI gene. Of the patients who presented with VEO-IBD (n=68), a diagnostic result was only achieved for three patients (4.4%). Excluding patients with VEO-IBD (n=68) increased the diagnostic yield to 33.9% (n=86/254 patients).

The most frequently implicated genes were *BTK* (n=5), *CD40LG* (n=5), *STAT3* (n=4), *NCF1* (n=4), and *CTLA4* (n=4). ‘Combined immunodeficiencies (CID) with associated or syndromic features’ was the most frequent IUIS diagnostic category (n=15) (Figure 4). Of the 48 implicated IUIS genes, 26 (54.2%) were autosomal recessive, one (2.1%) was autosomal recessive with increased susceptibility, eight (16.7%) were autosomal dominant, five (10.4%) autosomal dominant gain-of-function (GOF), two (4.2%) autosomal dominant loss-of-function (LOF), one (2.1%) was autosomal dominant with a dominant-negative effect and five (10.4%) were X-linked. Notably, *STAT3* variants were represented in two IUIS categories reflecting both autosomal dominant GOF and LOF mechanisms.

**Figure 4:**
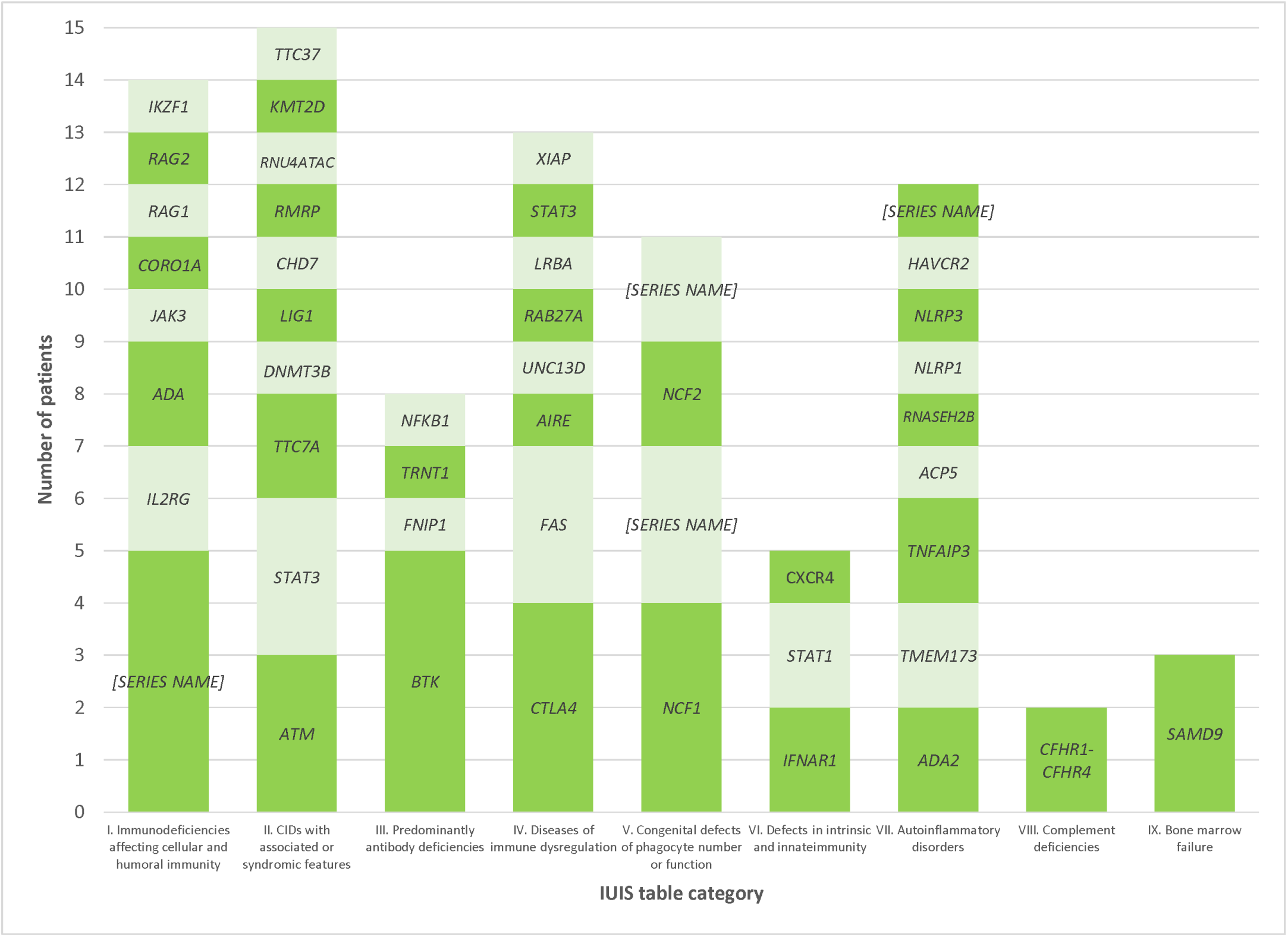
Implicated IUIS Genes for Diagnostic Cases.

Of the 446 unique instances of genomic testing, 111 tests (24.9%) yielded a VUS and 14 (3.1%) identified a risk allele associated with increased risk for Crohn’s disease and common variable immunodeficiency (CVID) (e.g., *NOD2* and *TACI*). Four of the 36 patients referred to genomic research received a positive result: two enrolled in research as a first-line approach (pre-MoC), with clinical testing confirming pathogenic variants in *CXCR4* and *CTLA4*, and two who underwent functional studies to assist with the classification of VUS in *CTLA4*,^20^ and *DOCK8*.^21^

Of the 10 (3.1%) deceased patients, six (60%) received a molecular diagnosis. Implicated genes included *RAG1, CYBB, ADA2, G6PC3, NCF2, CHD7,* or *SAMD9*. Causes of death included complications following HSCT (n=5) or sequelae of disease (n=5).

### Patient Management Outcomes

Management change recommendations informed by molecular diagnosis were investigated for patients included in the IUIS table (n=84) (**Supplementary Table 2**). A management change was recommended for 90.5% (n=76/84) of patients. A major management change recommendation occurred for 69.1% of patients (n=58/84), with ten patients receiving two forms of major management changes (n=68 total major treatment changes). Changes included recommendation for HSCT (n=29), immunoglobulin replacement therapy (IRT) (n=4), prophylaxis monitoring (n=7), selection of conditioning regimen (n=1), and other targeted treatment or management (n=27). A minor management change occurred for 21.4% of patients (n=18/84), all of which were changes to existing monitoring (n=15). Variations in management recommendations were observed among individuals with the same impacted gene and within familial cases, reflecting phenotypic variability and the timing of genomic testing. For instance, HSCT was initiated earlier in relatives due to a known familial variant, or a genomic diagnosis was established after targeted treatment had been delivered. No management change was recorded for 9.5% of patients (n=8/84), as no change occurred or data could not be identified.

### Impact of Model-of-care

All genomic tests that were excluded from the study due to results not being verifiable (n=31) were ordered pre-formal establishment of the MoC (<2023). Similarly, all 35 instances of duplicate testing occurred pre-Moc (<2021) (n=35/251;13.9%), compared to no duplicate testing post-MoC (≥2021) (n=0/230;0%) (p>0.05, chi-square test).

Significant improvements in genomic testing practices were noted for QPIAS initiated genomic testing (n=249/481), including increased diagnostic yield over time. Specifically, pre-MoC implementation (<2018), 45 genomic tests were ordered with a diagnostic yield of 26.1% (n=12/45) (Figure 5). There was a substantial increase in genomic testing post introduction of point-of-care genomic testing (2019-2020), which also correlated with a reduction in diagnostic yield (n=7/71, 9.9%). Post-MoC implementation (≥2021), the number of genomic tests ordered reduced, with the service delivering an average of 23 genomic tests per year and an average diagnostic rate of 27.4%. Overall, diagnostic yield significantly improved over time, increasing from 16.2% (n=19/117) pre-MoC (<2021) to 27.4% (n=32/117) post-MoC (≥2021) (p<0.05, chi-squared). Changes in testing laboratories were also noted, with 16.6% (n=18/108) of requested (excluding research and familial testing) conducted by Pathology Queensland pre-MoC (<2021), compared to 87.0% (n=100/115) post-MoC (≥2021) (**Supplementary figure 1**).

**Figure 5:**
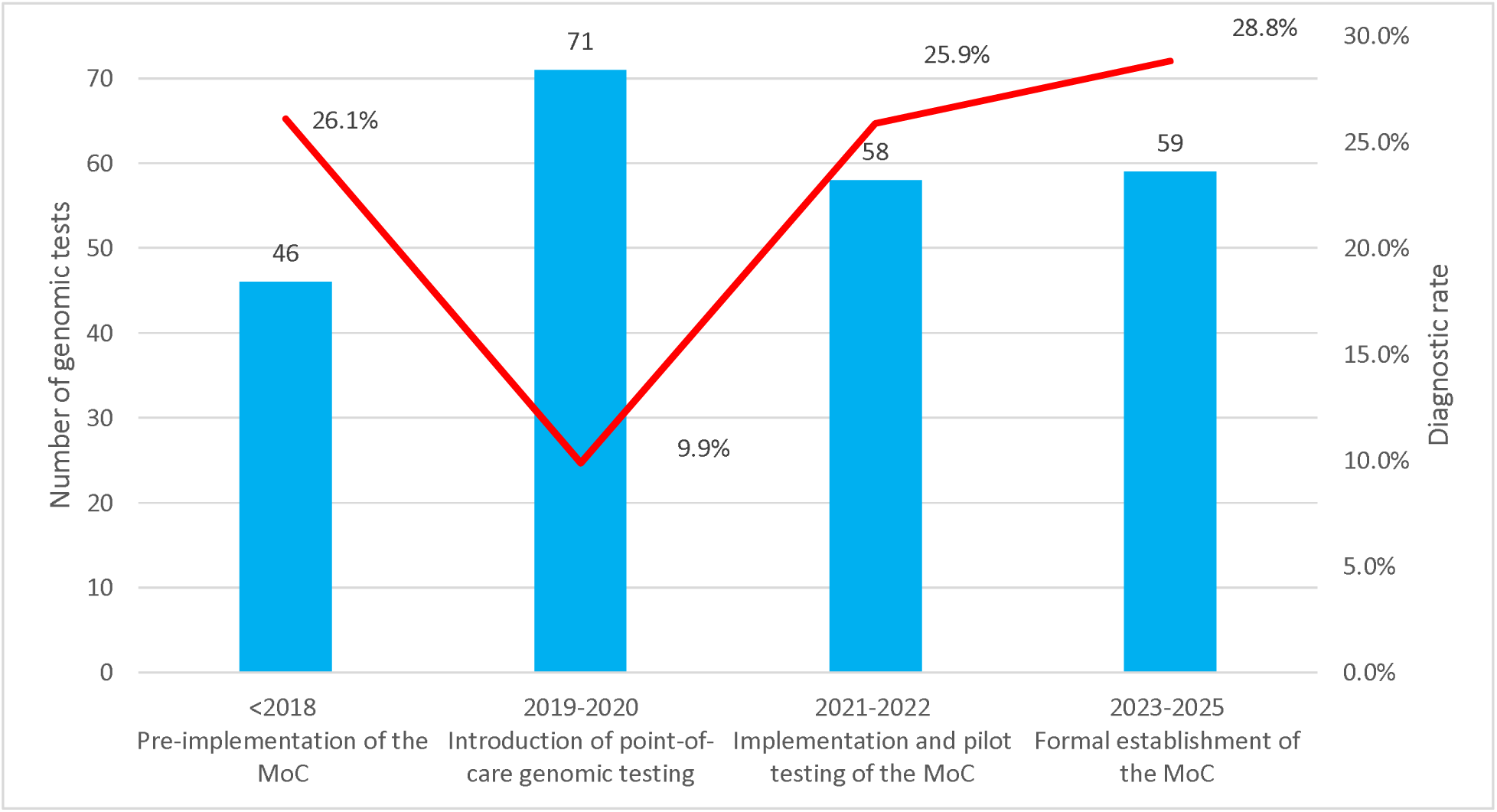
Changes in diagnostic genomic test (n=234) ordered by QPIAS immunologists compared to diagnostic rate over time*. *Excluded familial testing; MoC: model-of-care

Documentation of genomic consent increased over time (**Supplementary Figure 2**). The rate of documented informed consent for immunology-initiated testing pre-MoC implementation (<2018) was 50%, which significantly improved to 84% in 2019-2020, and 94% in 2023-2025. Overall, consent documentation significantly improved from 70.5% pre-MoC (<2021) to 88.4% post-MoC (≥2021). Additionally, there were changes to rates of VUS reported in testing initiated via QPIAS (inclusive of duplicate testing), pre-MoC implementation (<2021) (n=46/122;37.7%) as compared to post-MoC (≥2021) (n=9/126;7.1%) (p<0.5). All three patients who received VUS in 2025 have been reviewed at the MDT and consented to research aimed at reclassifying these results.

## Discussion

This study characterizes a pediatric IEI cohort and demonstrates the clinical and service effectiveness of implementing a genomic MoC. Our diagnostic yield of 27.6% is comparable to other IEI studies (average yield: 29%, range 10-70%).^22^ However, the diagnostic rate varied over time, improving alongside the implementation of the MoC. Management changes occurred for most patients with a diagnostic result.

Building on our previous feasibility assessment and parental evaluation of the MoC,^11,23^ this study identified improvements in genomic testing practices, such as test selection and documentation, and eliminated duplicate testing. Thus, providing further evidence to the effectiveness of establishing MoC to optimally mainstream in pediatric healthcare.

In keeping with international cohorts,^3,24,25^ CID with associated or syndromic features was the most common molecular diagnosis. The diagnostic rate in this study was highest among patients with phagocytic defects, SCID, and syndromic presentations, which is consistent with findings from other IEI studies.^25–27^ Conversely, the diagnostic rate was lowest among patients with VEO-IBD, a complex condition associated with IBD presenting under age six years.^28,29^ Monogenic etiology of IBD is associated with a younger age-of-onset, and a more aggressive and treatment-refractory disease course. Our low diagnostic yield for VEO-IBD is likely reflective of initial broad eligibility criteria that included offering genomic testing to all children diagnosed <6 years-of-age, regardless of additional phenotypic features. Of the three patients with a confirmed molecular diagnosis for VEO-IBD in our cohort, all were diagnosed under two years-of-age and presented with severe symptoms (e.g., congenital diarrhea, suspicious of VEO-Behcet’s disease, and infantile enterocolitis). Revised eligibility criterion was implemented in 2025 to automatically offer genomic testing to those with neonatal- and infantile-onset IBD (i.e., <2 years of age), and review VEO-IBD diagnosed over two years for additional features of monogenic IBD and IEI, including immunophenotyping, family history and disease severity. This adaptive approach to our MoC is another key strength of the study, whereby active monitoring of genomic data can directly inform clinical practice and local service processes.

Our findings demonstrate the importance of ongoing investigations of non-diagnostic results, with 20% of diagnoses occurring in subsequent testing, including research studies, expanded panels, and alternative testing modalities. Re-analysis of genomic data was limited to four patients within our cohort, which likely reflects the short timeframe of the MoC implementation, limiting opportunities for re-analysis at the recommended 2–3 year interval.^30^ Given the rapid rate of gene discovery for IEI, genomic test re-analysis will play an important role in enhancing diagnostic yields and should be considered for future genomic MoC.^31,32^

Our study adds to the evidence on the utility of genomic testing for IEI management, with 90.5% of patients with a molecular diagnosis in this cohort receiving a management change.^7^ While genomic testing can alter IEI treatment, it is important that it is delivered early in the disease course to mitigate irreversible disease related complications and use contraindicated treatments. For example, HSCT requires conditioning treatment via chemotherapy to prepare for transplant. However, conditioning therapy should consider molecular basis of disease to prevent toxicity (e.g. *LIG1* radiosensitive SCID).^33^ Other therapeutic changes can include initiation of targeted treatment, such as commencing JAK inhibitors for *STAT1* and *STAT3* gain-of-function variants,^34^ anti-TNF inhibitors for *RelA*-related autoinflammatory disease,^35^ PI3K-delta inhibitors for *APDS*,^36^ and abtacept for treatment refractory CTLA4-haploinsufficiency.^37^ Despite the impact of molecular diagnoses on IEI treatment, there is no Australian Medicare Item Number to facilitate testing, which currently offers testing for 25 other conditions (e.g., pediatric intellectual disability, cardiovascular disease, metabolic conditions).^38^ Public testing costs for IEI are instead covered by individual hospitals and health services, potentially resulting in access disparities across geographical locations.^39^ However, the availability of funding schemes alone does not guarantee uptake, as evidenced by the low use of existing Medicare rebates.^40^ Therefore, it is important to develop implementation strategies to support the adoption of genomic testing, including an adequate workforce, genomic education, and MoC that incorporate genomic considerations.

This study demonstrates the importance and efficiency of implementing a genomic MoC to address the complexities of delivering pediatric IEI genomic testing. While the study design did not permit association analysis to evaluate the impact of MoC on genomic testing practices, it is reasonable to infer certain patterns from the findings. The improved rate of consent is likely indicative of practice changes, including scheduled pre-test genetic counselling appointments and a new process for electronic filling of consent forms. The decrease in VUS rate is suggested to reflect strengthened collaboration with the local pathology service, who attend the monthly immunogenetic MDT meetings, conduct variant prioritization meetings with clinicians, and has facilitated most IEI genomic testing post-MoC. Implementation of MDT case review likely reduced duplicate testing and improved patient selection.

The importance of detailed clinical characterization was demonstrated in this study, substantially impacting diagnostic yield over time. Nevertheless, areas for improvement were identified, including the need for greater across specialty collaboration, given that over half of genomic tests in this cohort were ordered by a non-immunology specialist.^41^ These findings are consistent with previous studies, which reported an average of 7.9 specialists involved in IEI care.^3^ Based on our findings, recommendations for delivering mainstream IEI genomic testing are detailed in **Box 1**.

#### Considerations for implementing a genomic MoC in pediatric IEI

1. Include service review processes to improve clinical guidelines, such as eligibility for genomic tests.
2. Establish an MDT approach (e.g., clinical immunologists, nurses, genetic healthcare providers, immunopathologists, genetic pathologists and other medical specialists involved in patient care) to improve test processes and provide holistic family care.
3. Ensure appropriate process for pre-test consent that support patient decisionmaking and appropriate documentation, noting emerging technologies to streamline such processes.^42,43^
4. Where possible, utilize trio-exome or genome sequencing to improve diagnostic yield and optimize health service resources.
5. For those with diagnostic results, consider the broader implications, including familial and reproductive considerations.
6. Increased collaboration with research entities and testing laboratories to resolve uncertain results or further investigate non-diagnostic results.^44^
7. Establishing processes for review and re-analysis following a negative or uncertain genomic finding

Findings should be interpreted in line with the limitations of this study. The retrospective design of the study required reliance on data primarily collected for clinical purposes, which is less accurate than data gathered prospectively. Data on the genetic counselling and reproductive implications of molecular diagnoses could not be comprehensively collected, as related decisions frequently occurred within other clinical services and extended beyond the patient whose chart was reviewed. Additionally, while the cohort was geographically and socio-economically representative of the wider Queensland population, significant inequities in access to genomic testing for pediatric IEI remain,^39^ such as those related to ancestral background, which were not captured in this study. Nevertheless, a key strength of the study is the description of genomic test utilization that captures real-world clinical practice over time. While a formal economic evaluation has not been undertaken, it is anticipated that these outcomes will be associated with cost savings for the public healthcare system. The comprehensive chart review and audit data allowed for IEI cohort and genomic test description, with findings further informing service improvement. With increasing clinical and personal utility of genomic testing in pediatric IEI and other specialties, there is a need to develop evidence based on MoC to maximize such outcomes. Our study demonstrates that integrating a targeted MoC that considers the complexities of genomic testing can result in substantial improvements in genomic testing practices, patient care, and service delivery.

## Data availability

De-identified data is available upon request to the corresponding author.

