## Supplementary Figures for "Evaluating mainstreaming in pediatric immunology: an optimal model of care"

### Supplementary Figure 1: Testing Laboratory

Genomic tests

|  | Domestic private | International | Other domestic public | Pathology Queensland |
| --- | --- | --- | --- | --- |
| <2018 Pre-implementation of the MoC | 11 | 25 | 4 | 1 |
| 2019-2020 Introduction of point-of-care genomic testing | 8 | 41 | 1 | 17 |
| 2021-2022 Implementation and pilot testing of the MoC | 1 | 8 | 1 | 46 |
| 2023-2025 Formal establishment of the MoC | 3 | 1 | 1 | 54 |

*Includes genomic testing initiated by immunology specialist. Excludes research and familial testing

MoC: model-of-care

### Supplementary Figure 2: Genomic Consent Documentation

|  | Consent documentation |
| --- | --- |
| <2018 Pre-implementation of the MoC | 50% |
| 2019-2020 Introduction of point-of-care genomic testing | 83.7838% |
| 2020-2021 Implementation and pilot testing of the MoC | 82.8125% |
| 2023-2025 Formal establishment of the MoC | 93.8462% |

MoC: model-of-care
