## Supplementary tables for "Evaluating mainstreaming in pediatric immunology: an optimal model of care"

### Supplementary Table 1: Chart review data

| **Item** | **Description** |
| --- | --- |
| Demographic information | Patient age, living status, biological sex, and postcode |
| Instances of genomic testing | Each instance of testing per patient, including duplications of testing |
| Genomic testing details | Date of genomic testing, ordering clinician and department, laboratory, testing modality and panels applied, and evidence of informed consent documentation |
| Genomic testing result | Date of result reported, result type (e.g., diagnostic), gene and variant details |
| Outcomes of genomic testing | Impact of genomic testing on treatment, management, reproductive planning, and referral to research pathway |

### Supplementary Table 2: Diagnostic results according to IUIS

| **ID** | **Gene** | **Variant details** | **Pathogenicity** | **Variant type** | **Zygosity** | **Condition** | **IUIS subclass** | **Inheritance** | **Major management change** | **Minor management change** |
| --- | --- | --- | --- | --- | --- | --- | --- | --- | --- | --- |
| **Table I: Immunodeficiencies affecting cellular and humoral immunity** | | | | | | | | | | |
| 1.1.1 | *IL2RG* | NM_000206.3:c.670C>T | PV | Missense | Hemizygous | gc deficiency (common gamma chain SCID, CD132 deficiency) | 1.T-B+ SCID | XL | Yes: HSCT | No |
| 1.1.2 | *IL2RG* | NM_000206.3:c.670C>T | PV | Missense | Hemizygous | gc deficiency (common gamma chain SCID, CD132 deficiency) | 1.T-B+ SCID | XL | Yes: HSCT | No |
| 1.2 | *CORO1A* | NM_007074.4: arr[GRCh37] 16p11.2(29614976_30215621)x1 | PV | Deletion | Heterozygous | Coronin-1A deficiency | 1.T-B+ SCID | AR | Yes: IRT and prophylaxis monitoring | No |
|  | *CORO1A* | NM_007074.4:c.919C>T | LPV | Missense | Heterozygous | Coronin-1A deficiency | 1.T-B+ SCID | AR |  |  |
| 1.3 | *JAK3* | NM_000215.4:c.2281del | PV | Not reported | Homozygous | JAK3 deficiency | 1.T-B+ SCID | AR | Yes: HSCT | No |
| 1.4 | *ADA* | NM_000022.4:c.872C>T | PV | Missense | Heterozygous | ADA deficiency SCID | 2.T-B-SCID | AR | Yes: Targeted treatment and management and HSCT | No |
|  | *ADA* | NM_000022.2:c.(?_-128)_(*312_?)del | PV | Deletion | Heterozygous | ADA deficiency SCID | 2.T-B-SCID | AR |  |  |
| 1.5 | *ADA* | NM_000022.4:c.478+1G>A | PV | Splice | Homozygous | ADA deficiency SCID | 2.T-B-SCID | AR | Yes: Targeted treatment and management and HSCT | No |
| 1.6 | *RAG1* | NM_000448.3:c.322C>T | PV | Not reported | Homozygous | RAG deficiency | 2.T-B-SCID | AR | No | No |
| 1.7 | *RAG2* | NM_000536.4:c.686G>A | PV | Missense | Heterozygous | RAG deficiency | 2.T-B- SCID | AR | Yes: HSCT | No |
|  | *RAG2* | NM_000536.4:c.859del | PV | Frameshift | Heterozygous | RAG deficiency | 2.T-B-SCID | AR |  |  |
| 1.8 | *CD40LG* | NM_000074.3:c.368C>A | PV | Missense | Hemizygous | CD40 ligand (CD154) deficiency | 3.CID, Generally Less Profound than SCID | XL | Yes: HSCT | No |
| 1.9 | *CD40LG* | NM_000074.3:c.478C>T | PV | Nonsense | Hemizygous | CD40 ligand (CD154) deficiency | 3.CID, Generally Less Profound than SCID | XL | Yes: HSCT | No |
| 1.10 | *CD40LG* | NM_000074.3:c.224dup | PV | Frameshift | Hemizygous | CD40 ligand (CD154) deficiency | 3.CID, Generally Less Profound than SCID | XL | Yes: HSCT | No |
| 1.11.1 | *CD40LG* | NM_000074.3:c.506_507del | PV | Frameshift | Hemizygous | CD40 ligand (CD154) deficiency | 3.CID, Generally Less Profound than SCID | XL | Yes: IRT | No |
| 1.11.2 | *CD40LG* | NM_000074.3:c.506_507del | PV | Frameshift | Hemizygous | CD40 ligand (CD154) deficiency | 3.CID, Generally Less Profound than SCID | XL | Yes: IRT | No |
| 1.12 | *IKZF1* | NM_006060.6:c.476A>G | PV | Missense | Heterozygous | IKAROS deficiency | 3.CID, Generally Less Profound than SCID | AD DN | Yes: HSCT | No |
| **Table II: Combined immunodeficiencies with associated or syndromic features** | | | | | | | | | | |
| 2.1 | *ATM* | NM_000051.4:c.1401dup | PV | Frameshift | Heterozygous | Ataxia-telangiectasia | 2.DNA Repair Defects | AR | No | Yes: Changes in monitoring |
|  | *ATM* | NM_000051.4:c.3802del | PV | Nonsense | Heterozygous | Ataxia-telangiectasia | 2.DNA Repair Defects | AR |  |  |
| 2.2 | *ATM* | NM_000051.4:c.3310del | PV | Frameshift | Homozygous | Ataxia-telangiectasia | 2.DNA Repair Defects | AR | No | Yes: Changes in monitoring |
| 2.3 | *ATM* | NM_000051.4:c.1564_1565del | PV | Frameshift | Heterozygous | Ataxia-telangiectasia | 2.DNA Repair Defects | AR | No | Yes: Changes in monitoring |
|  | *ATM* | NM_000051.4:c.3802del | PV | Nonsense | Heterozygous | Ataxia-telangiectasia | 2.DNA Repair Defects | AR |  |  |
| 2.4 | *DNMT3B* | NM_006892.4:c.2506G>C | PV | Missense | Homozygous | Immunodeficiency with centromeric instability and facial anomalies type 1 | 2.DNA Repair Defects | AR | No | Yes: Changes in monitoring |
| 2.5 | *LIG1* | NM_000234.3:c.1244del | PV | Splice | Heterozygous | Ligase I deficiency | 2.DNA Repair Defects | AR | Yes: Changes to conditioning regimen | No |
|  | *LIG1* | NM_000234.3:c.2005-1107G>A | PV | Frameshift | Heterozygous | Ligase I deficiency | 2.DNA Repair Defects | AR |  |  |
| 2.6 | *CHD7* | NM_017780.4:c.5932del | PV | Frameshift | Heterozygous | CHARGE syndrome | 3.Thymic Defects with Additional Congenital Anomalies | AD | No | No |
| 2.7 | *RMRP* | NC_000009.11:g.35657945T>C | PV | Intronic | Homozygous | Cartilage hair hypoplasia | 4.Immuno-osseous Dysplasias | AR | No | Yes: Changes in monitoring |
| 2.8 | *RNU4ATAC* | NC_000002.11:g.122288572A>G | PV | Intronic | Heterozygous | MOPD1 Deficiency (Roifman syndrome) | 4.Immuno-osseous Dysplasias | AR | No | Yes: Changes in monitoring |
|  | *RNU4ATAC* | NC_000002.11:g.122288468C>T | PV | Intronic | Heterozygous | MOPD1 Deficiency (Roifman syndrome) | 4.Immuno-osseous Dysplasias | AR |  |  |
| 2.9 | *STAT3* | NM_139276.3:c.1780G>A | LPV | Missense | Heterozygous | AD-HIES STAT3 deficiency (Job syndrome) | 5.Hyper IgE Syndromes | AD LOF (DN) | Yes: Targeted treatment and management | No |
| 2.10 | *STAT3* | NM_139276.3:c.1897C>T | LPV | nonsense | Heterozygous | AD-HIES STAT3 deficiency (Job syndrome) | 5.Hyper IgE Syndromes | AD LOF (DN) | Yes: Targeted treatment and management | No |
| 2.11 | *STAT3* | NM_139276.3: c.1234A>G | LPV | Missense | Heterozygous | AD-HIES STAT3 deficiency (Job syndrome) | 5.Hyper IgE Syndromes | AD LOF (DN) | Yes: Targeted treatment and management and HSCT | No |
| 2.12 | *KMT2D* | NM_003482.3:c.(?_1)(2797+1_2798-1)del | LPV | Deletion | Heterozygous | Kabuki Syndrome type 1 | 9.Other Defects | AD | No | Yes: Changes in monitoring |
| 2.13 | *TTC37 (SKIC3)* | NM_014639.3:c.1307_1308del | Not reported | Not reported | Heterozygous | Tricho-Hepato-Enteric Syndrome | 9.Other Defects | AR | Yes: IRT | No |
|  | *TTC37 (SKIC3)* | NM_014639.3:c.1374C>G | Not reported | Nonsense | Heterozygous | Tricho-Hepato-Enteric Syndrome | 9.Other Defects | AR |  |  |
| 2.14.1 | *TTC7A* | NM_020458.4:c.2225-2A>G | LPV | Splice | Heterozygous | Gastrointestinal defects and immunodeficiency syndrome | 9.Other Defects | AR | No | No |
|  | *TTC7A* | NM_020458.4:c.517+1G>C | LPV | Splice | Heterozygous | Gastrointestinal defects and immunodeficiency syndrome | 9.Other Defects | AR |  |  |
| 2.14.2 | *TTC7A* | NM_020458.4:c.2225-2A>G | LPV | Splice | Heterozygous | Gastrointestinal defects and immunodeficiency syndrome | 9.Other Defects | AR | No | No |
|  | *TTC7A* | NM_020458.4:c.517+1G>C | LPV | Splice | Heterozygous | Gastrointestinal defects and immunodeficiency syndrome | 9.Other Defects | AR |  |  |
| **Table III: Predominantly Antibody Deficiencies** | | | | | | | | | | |
| 3.1 | *BTK* | NM_000061.3:c.34A>C | LPV | Missense | Hemizygous | BTK deficiency, XLA | 1. Severe Reduction in All Serum Immunoglobulin Isotypes with Profoundly Decreased or Absent B Cells, Agammaglobulinemia | XL | No | Yes: Changes in monitoring |
| 3.2 | *BTK* | NM_000061.3:c.904G>A | LPV | Missense | Hemizygous | BTK deficiency, XLA | 1. Severe Reduction in All Serum Immunoglobulin Isotypes with Profoundly Decreased or Absent B Cells, Agammaglobulinemia | XL | No | Yes: Changes in monitoring |
| 3.3 | *BTK* | NM_000061.3:c.1349+5G>A | PV | Not reported | Hemizygous | BTK deficiency, XLA | 1. Severe Reduction in All Serum Immunoglobulin Isotypes with Profoundly Decreased or Absent B Cells, Agammaglobulinemia | XL | No | Yes: Changes in monitoring |
| 3.4 | *BTK* | NM_000061.3:c.215dup | PV | Frameshift | Hemizygous | BTK deficiency, XLA | 1. Severe Reduction in All Serum Immunoglobulin Isotypes with Profoundly Decreased or Absent B Cells, Agammaglobulinemia | XL | No | Yes: Changes in monitoring |
| 3.5 | *BTK* | NM_000061.3:c.136dup | Not reported | Not reported | Hemizygous | BTK deficiency, XLA | 1. Severe Reduction in All Serum Immunoglobulin Isotypes with Profoundly Decreased or Absent B Cells, Agammaglobulinemia | XL | No | Yes: Changes in monitoring |
| 3.6 | *FNIP1* | NM_133372.3:c.2773dup | PV | Frameshift | Homozygous | FNIP1 deficiency | 1.Severe Reduction in All Serum Immunoglobulin Isotypes with Profoundly Decreased or Absent B Cells, Agammaglobulinemia | AR | No | Yes: Changes in monitoring |
| 3.7 | *NFKB1* | NM_003998.4:c.448del | LPV | Frameshift | Heterozygous | NFKB1 deficiency | 2.Severe Reduction in at Least 2 Serum Immunoglobulin Isotypes with Normal or Low Number of B Cells, CVID Phenotype | AD | No | Yes: Changes in monitoring |
| 3.8 | *TRNT1* | NM_182916.3:c.1246A>G | LPV | Missense | Heterozygous | TRNT1 deficiency | 2.Severe Reduction in at Least 2 Serum Immunoglobulin Isotypes with Normal or Low Number of B Cells, CVID Phenotype | AR | No | No |
|  | *TRNT1* | NM_182916.3:c.803-2A>G | LPV | Splice | Heterozygous | TRNT1 deficiency | 2.Severe Reduction in at Least 2 Serum Immunoglobulin Isotypes with Normal or Low Number of B Cells, CVID Phenotype | AR |  |  |
| **Table IV: Diseases of Immune Dysregulation** | | | | | | | | | | |
| 4.1 | *UNC13D* | NM_199242.3:c.118-308C>T | LPV | Missense | Heterozygous | UNC13D / Munc13-4 deficiency (FHL3) | 1. Familial Hemophagocytic Lymphohistiocytosis | AR | Yes: HSCT | No |
|  | *UNC13D* | NM_199242.3:c.1820G>C | LPV^ | Missense | Heterozygous | UNC13D / Munc13-4 deficiency (FHL3) | 1. Familial Hemophagocytic Lymphohistiocytosis | AR |  |  |
| 4.2 | *RAB27A* | NM_004580.4:c.344-?467+?del | PV | Deletion | Homozygous | Griscelli syndrome, type 2 | 2. FHL Syndromes with Hypopigmentation | AR | Yes: HSCT | No |
| 4.3.1 | *CTLA4* | NM_005214.5:c.118G>A | LPV | Missense | Heterozygous | CTLA4 haploinsufficiency (ALPS-V) | 3.Regulatory T Cell Defects | AD | No | No |
| 4.3.2 | *CTLA4* | NM_005214.5:c.118G>A | LPV | Missense | Heterozygous | CTLA4 haploinsufficiency (ALPS-V) | 3.Regulatory T Cell Defects | AD | Yes: Targeted treatment and management | No |
| 4.3.3 | *CTLA4* | NM_005214.5:c.118G>A | LPV | Missense | Heterozygous | CTLA4 haploinsufficiency (ALPS-V) | 3.Regulatory T Cell Defects | AD | Yes: Targeted treatment and management and HSCT | No |
| 4.3.4 | *CTLA4* | NM_005214.5:c.118G>A | LPV | Missense | Heterozygous | CTLA4 haploinsufficiency (ALPS-V) | 3.Regulatory T Cell Defects | AD | Yes: Targeted treatment and management and HSCT | No |
| 4.4 | *LRBA* | NM_001364905.1:c.4759_4762del | PV | Frameshift | Heterozygous | LRBA deficiency | 3.Regulatory T Cell Defects | AR | Yes: Targeted treatment and management and HSCT | No |
|  | *LRBA* | NM_001364905.1: c.7843_7846del | PV | Frameshift | Heterozygous | LRBA deficiency | 3.Regulatory T Cell Defects | AR |  |  |
| 4.5 | *STAT3* | NM_139276.3:c.737G>A | LPV | Missense | Heterozygous | STAT3 GOF | 3.Regulatory T Cell Defects | AD GOF | Yes: Targeted treatment and management | No |
| 4.6 | *AIRE* | NM_000383.4:c.967_979 del | PV | Frameshift | Homozygous | APECED (APS-1), autoimmune polyendocrinopathy with candidiasis and ectodermal dystrophy | 4.Autoimmunity with or without Lymphoproliferation | AR | No | No |
| 4.7 | *FAS* | NM_000043.6:c.657_658del | LPV | Frameshift | Heterozygous | Apoptosis defect FAS mediated | 6.Autoimmune Lymphoproliferative Syndrome | AD | Yes: Targeted treatment and management | No |
| 4.8.1 | *FAS* | NM_000043.6:c.761T>G | LPV | Missense | Heterozygous | Apoptosis defect FAS mediated | 6.Autoimmune Lymphoproliferative Syndrome | AD | Yes: Targeted treatment and management | No |
| 4.8.2 | *FAS* | NM_000043.6:c.761T>G | LPV | Missense | Heterozygous | Apoptosis defect FAS mediated | 6.Autoimmune Lymphoproliferative Syndrome | AD | Yes: Targeted treatment and management | No |
| 4.9 | *XIAP* | NM_001167.4:c.1048G>T | PV | Nonsense | Hemizygous | Lymphoproliferative Syndrome, 2 | 7.Susceptibility to EBV and Lymphoproliferative Conditions | XL | Yes: HSCT | No |
| **Table V: Congenital defects of phagocyte number or function** | | | | | | | | | | |
| 5.1.1 | *ITGB2* | NM_000211.5:c.532C>T | LPV | Missense | Homozygous | LAD1 | 2.Defects of Motility | AR | Yes: Targeted treatment and management and HSCT | No |
| 5.1.2 | *ITGB2* | NM_000211.5:c.532C>T | LPV | Missense | Homozygous | LAD1 | 2.Defects of Motility | AR | Yes: Targeted treatment and management and HSCT | No |
| 5.1.3 | *ITGB2* | NM_000211.5:c.532C>T | LPV | Missense | Homozygous | LAD1 | 2. Defects of Motility | AR | Yes: Targeted treatment and management and HSCT | No |
| 5.2 | *NCF1* | NM_000265.7:c.604C>T | LPV | nonsense | Heterozygous | AR CGD | 3.Defects of Respiratory Burst | AR | Yes: HSCT | No |
|  | *NCF1* | NM_000265.7:c.923C>T | PV | Missense | Heterozygous | AR CGD | 3.Defects of Respiratory Burst | AR |  |  |
| 5.3.1^^ | *NCF1* | NM_000265.7:c.75_76del | PV | Frameshift | Homozygous | AR CGD | 3.Defects of Respiratory Burst | AR | Yes: Prophylaxis monitoring | No |
| 5.3.2^^ | *NCF1* | NM_000265.7:c.75_76del | PV | Frameshift | Homozygous | AR CGD | 3.Defects of Respiratory Burst | AR | Yes: Prophylaxis monitoring | No |
| 5.4 | *NCF1* | NM_000265.7:c.75_76del | PV | Frameshift | Homozygous | AR CGD | 3.Defects of Respiratory Burst | AR | Yes: HSCT | No |
| 5.5 | *NCF2* | NM_000433.4:c.482del | PV | Frameshift | Homozygous | AR CGD | 3.Defects of Respiratory Burst | AR | Yes: HSCT | No |
| 5.6 | *NCF2* | NM_000433.4:c.482del | PV | Frameshift | Homozygous | AR CGD | 3.Defects of Respiratory Burst | AR | Yes: HSCT | No |
| 5.7.1 | *CYBB* | NM_000397.4:c.(674+1_675-1)_(804+1_805-1)del | PV | Complex structural variant | Hemizygous | XL CGD gp91phox | 3.Defects of Respiratory Burst | XL | Yes: HSCT | No |
| 5.7.2 | *CYBB* | NM_000397.4:c.(674+1_675-1)_(804+1_805-1)del | PV | Complex structural variant | Hemizygous | XLCGD, gp91phox | 3.Defects of Respiratory Burst | XL | Yes: HSCT | No |
| **Table VI: Defects in Intrinsic and Immunity** | | | | | | | | | | |
| 6.1.1 | *STAT1* | NM_007315.4:c.800C>T | PV | Missense | Heterozygous | STAT1 deficiency | 1. MSMD | AD LOF | Yes: Prophylaxis monitoring | No |
| 6.1.2 | *STAT1* | NM_007315.4:c.800C>T | PV | Missense | Heterozygous | STAT1 deficiency | 1.MSMD | AD LOF | Yes: Prophylaxis monitoring | No |
| 6.2 | *CXCR4* | NM_003467.3:c.1000C>T | PV | Nonsense | Heterozygous | WHIM syndrome | 2.Epidermodysplasia verruciformis (HPV) | AD GOF | Yes: Targeted treatment and management | No |
| 6.3 | *IFNAR1* | NM_000629.3:c.1156G>T | PV | Nonsense | Homozygous | IFNAR1 deficiency | 3.Predisposition to Severe Viral Infection | AR | Yes: Prophylaxis monitoring | No |
| 6.4 | *IFNAR1* | NM_000629.3:c.1156G>T | PV | Nonsense | Homozygous | IFNAR1 deficiency | 3.Predisposition to Severe Viral Infection | AR | Yes: Prophylaxis monitoring | No |
| **Table VII: Autoinflammatory Disorders** | | | | | | | | | | |
| 7.1 | *ACP5* | NM_001611.5:c.131C>T | LPV | Missense | Heterozygous | SPENCD | 1.Type 1 Interferonopathies | AR | No | Yes: Changes in monitoring |
|  | *ACP5* | NM_001611.5:c.712T>C | PV | Missense | Heterozygous | SPENCD | 1.Type 1 Interferonopathies | AR |  |  |
| 7.2.1 | *ADA2* | NM_001282225.2:c.506G>A | LPV | Missense | Homozygous | ADA2 deficiency | 1.Type 1 Interferonopathies | AR | Yes: Targeted treatment and management | No |
| 7.2.2 | *ADA2* | NM_001282225.2:c.506G>A | LPV | Missense | Homozygous | ADA2 deficiency | 1.Type 1 Interferonopathies | AR | Yes: Targeted treatment and management | No |
| 7.3 | *RNASEH2B* | NM_024570.4:c.529G>A | LPV | Missense | Homozygous | Aicardi-Goutiers Syndrome | 1.Type 1 Interferonopathies | AR | No | No |
| 7.4 | *TMEM173* | NM_198282.4:c.463G>A | PV | Missense | Heterozygous | STING-associated vasculopathy, infantileonset | 1.Type 1 Interferonopathies | AD | Yes: Targeted treatment and management | No |
| 7.5 | *TMEM173* | NM_198282.4:c.463G>A | PV | Missense | Heterozygous | STING-associated vasculopathy, infantileonset | 1.Type 1 Interferonopathies | AD | Yes: Targeted treatment and management | No |
| 7.6 | *NLRP1* | NM_033004.4:c.230T>C | PV | Missense | Heterozygous | Autoinflammation with arthritis and dyskeratosis | 2.Defects Affecting the Inflammasome | AD GOF | Yes: Targeted treatment and management | No |
| 7.8 | *NLRP3* | NM_001243133.2:c.1322C>T | PV | Missense | Heterozygous | Muckle-Wells syndrome | 2.Defects Affecting the Inflammasome | AD GOF | Yes: Targeted treatment and management | No |
| 7.9 | *HAVCR2* | NM_032782.5:c.245A>G | P | Missense | Homozygous | T-cell lymphoma subcutaneous panniculitislike (TIM3 deficiency) | 3.Non-Inflammasome Related Conditions | AR | Yes: HSCT | No |
| 7.10 | *TNFAIP3* | NM_001270508.2:c.486+1G>A | LPV | Splice | Heterozygous | Haploinsufficiency of A20/HA20 | 3.Non-Inflammasome Related Conditions | AD | Yes: HSCT | No |
| 7.11 | *TNFAIP3* | NM_001270508.2:c.486+1G>A | LPV | Splice | Heterozygous | Haploinsufficiency of A20/HA20 | 3.Non-Inflammasome Related Conditions | AD | Yes: Targeted treatment and management | No |
| 7.12 | *RELA* | NM_021975.4:c.1334_1335insG | LPV | Frameshift | Heterozygous | RELA haploinsufficiency/ interferonopathy | 1.Type 1 Interferonopathies | AD | Yes: Targeted treatment and management | No |
| **Table VIII: Complement Deficiencies** | | | | | | | | | | |
| 8.1 | *CFHR3-CFHR1* | NC_000001.10:g.196743970_196764537del  NC_000001.10:g.196788861_196801319del | Not reported | Deletion | Heterozygous | Factor H –related protein deficiencies | 1.Complement Deficiencies | AR (Increased susceptibility) | Yes: Targeted treatment and management | No |
|  | *CFHR1-CFHR4* | NC_000001.10:g.196788861_196801319del  NC_000001.10:g.196857182_196887763del | Not reported | Deletion | Heterozygous | Factor H–related protein deficiencies | 1.Complement Deficiencies | AR (Increased susceptibility) |  |  |
| 8.2 | *CFHR1-CFHR4* | *arr[GRCh37] 1q31.3(196801025_196892322)x0* | Not reported | Deletion | Homozygous | Factor H–related protein deficiencies | 1.Complement Deficiencies | AR (Increased susceptibility) | Yes: Targeted treatment and management | No |
| **Table IX: Bone Marrow Failure** | | | | | | | | | | |
| 9.1 | *SAMD9* | NM_017654.4:c.1868C>T | LPV | Not reported | Heterozygous | MIRAGE | 1.Bone Marrow Failure | AD GOF | No | Yes: Changes in monitoring |
| 9.2 | *SAMD9* | NM_017654.4:c.2318T>C | LPV | Missense | Heterozygous | MIRAGE | 1.Bone Marrow Failure | AD GOF | Yes: HSCT | Yes: Changes in monitoring |
| 9.3 | *SAMD9* | NM_017654.4:c.2318T>C | LPV | Missense | Heterozygous | MIRAGE | 1.Bone Marrow Failure | AD GOF | No | Yes: Changes in monitoring |

^ Pathogenicity upgraded from VUS by functional testing

^^ Methodology high likelihood via Southern Blot due to highly homologous pseudogenes in NCF1b and NCF1c

ADA: Adenosine deaminase; AD: Autosomal dominant; AR: Autosomal recessive; CGD: Chronic granulomatous disease; CID: Combined immunodeficiency; CVID: Common variable immunodeficiency; DN: Dominant negative; GOF: Gain of function; HSCT: Haematopoietic stem cell transplantation; IRT: Immunoglobulin replacement therapy; LAD1: Leukocyte adhesion deficiency type 1; LPV: Likely pathogenic variant; LOF: Loss of function; MIRAGE: Myelodysplasia, infection, restriction of growth, adrenal hypoplasia, genital phenotypes, enteropathy; MSMD: Mendelian susceptibility to mycobacterial disease; PV: Pathogenic variant; SCID: Severe combined immunodeficiency; SPENCD: Spondyloenchondrodysplasia with immune dysregulation; WHIM: Warts, hypogammaglobulinemia, infections, myelokathexis; XL: X-linked; XLA: X-linked agammaglobulinemia
